# Real world performance of inactivated SARS-CoV-2 vaccine (CoronaVac) against infection, hospitalization and death due to COVID-19 in adult population in Indonesia

**DOI:** 10.1101/2022.02.02.22270351

**Authors:** Anton Suryatma, Raras Anasi, Miko Hananto, Asep Hermawan, Ririn Ramadhany, Irene Lorinda Indalao, Agustiningsih Agustiningsih, Ely Hujjatul Fikriyah, Kristina Lumban Tobing, Teti Tejayanti, Rustika Rustika, Pandji Wibawa Dhewantara

**Affiliations:** National Agency for Research and Innovation, Jl. M.H. Thamrin No. 8, Jakarta 10340, Indonesia; Center for Policy on Global Health and Health Technology, Institute of Health Policy Development, Ministry of Health of Indonesia, Jl. Percetakan Negara No. 29, Jakarta 10560, Indonesia; Center for Policy on Health System and Resources, Institute of Health Policy Development, Ministry of Health of Indonesia, Jl. Percetakan Negara No. 29, Jakarta 10560, Indonesia

**Keywords:** inactivated vaccine, COVID-19, effectiveness, test-negative, Indonesia

## Abstract

**Background:** Inactivated SARS-CoV-2 vaccine has been included in the national COVID-19 vaccination program in Indonesia since January 2021. The study aims to assess the impacts of inactivated COVID-19 vaccine on infection, hospitalization, and death among adult population aged ≥18 years in Bali, Indonesia.

**Methods:** Test-negative, case control study was conducted by linking SARS-CoV-2 laboratory records, vaccination, and health administrative data for the period of January 13 to June 30, 2021. Case-subjects were defined as individuals who had a positive RT-PCR test for SARS-CoV-2 during the period; they were matched with controls (tested negative) (1:1) based on age, sex, district of residence, and week of testing. We estimated the odds of vaccination in PCR confirmed, hospitalization and death due to COVID-19, accounting for the presence of comorbidities and prior infection. Vaccine effectiveness was estimated as (1-odds ratio) x 100%.

**Results:** Total 109,050 RT-PCR test results were retrieved during the January 13 to June 30, 2021. Of these, 14,168 subjects were eligible for inclusion in the study. Total 5518 matched case-control pairs were analyzed. Adjusted vaccine effectiveness (VE) against laboratory-confirmed SARS-CoV-2 infection was 14.5% (95% confidence interval -11 to 34.2) at 0-13 days after the first dose; 66.7% (95% CI: 58.1-73.5) at ≥14 days after the second dose. The adjusted effectiveness against hospitalization and COVID-19-associated death was 71.1% (95% CI: 62.9-77.6) and 87.4% (95% CI: 65.1-95.4%) at ≥14 days after receiving the second dose, respectively.

**Conclusions:** Two-dose of inactivated CoronaVac vaccine showed high effectiveness against laboratory confirmed COVID-19 infection, hospitalization, and death associated with COVID-19 among adults aged ≥18 years.

## Introduction

Indonesia rolled-out the first phase of mass vaccination program on January 13th, 2021, using the inactivated SARS-CoV-2 vaccine, CoronaVac, manufactured by Sinovac Life Sciences, Beijing, China. In the first phase, mass COVID-19 vaccination was targeting healthcare workers and general population including adults and elderly. A phase 1 and 2 clinical trial in China indicated that CoronaVac was tolerable, with acceptable safety and immunogenicity, and therefore supported the conduct of a phase 3 clinical trial in three countries including Indonesia [1]. The efficacy of CoronaVac varied among the countries, ranging from 50.7% to 84% against SARS-CoV-2 infection [2-4]. In Indonesia, two doses of CoronaVac vaccine had an efficacy of 65.3% against COVID-19 illness after 14 days of injection; this supported the approval of emergency use authorization (EUA) by the Indonesia National Agency of Drug and Food Control (NADFC) [2, 5].

Evidence regarding the real-world effectiveness of inactivated CoronaVac vaccine against SARS-CoV-2 infection, hospitalization, and death related with COVID-19 have been reported in number of countries such as Chile, Turkey, and Brazil [6, 7]. These studies suggest that the effectiveness of CoronaVac was varied from region to region. Therefore, since the performance in the population found to be different from that reported efficacy under controlled trial circumstances, post-licensure vaccine effectiveness evaluation at the population level is crucial [8]. In Indonesia, study on evaluating CoronaVac vaccine performance in preventing COVID-19 infection and severe outcomes is still limited especially in adults aged 18 years and above. The objective of this study was to estimate vaccine effectiveness (VE) of inactivated CoronaVac vaccine against COVID-19 infection, hospitalization, and COVID-19-related death among adult aged ≥18 years in Bali.

## Materials and methods

### Setting and design

Bali is a one of the popular tropical islands in Indonesia, with a total land area of 5780 km^2^ and population of more than 4.3 million [9]. Mass COVID-19 vaccination using inactivated SARS-CoV-2 (CoronaVac) was initiated by the Ministry of Health of Indonesia on January 13, 2021, across Indonesia, including in Bali. As of July 31, 2021, COVID-19 vaccines have been given to 2.8 million (41.9%) of the targeted Balinese population. About 2.2 million people aged ≥18 years had received at least one dose of COVID-19 vaccines (Supplementary Figure S1); 1.2 million (43.1%) had received at least one dose of inactivated CoronaVac vaccine [10]. The observation was conducted during January 13 to June 30, 2021. During the study period, the incidence of COVID-19 was increased during January-February and followed with a declining trend from March to June 2021. There was a sharp increase in incidence in July due to B.1.617.2 (Delta) variants (Figure 2, Supplementary Figure S2). We conducted a record-based retrospective, test-negative, matched case control (1:1) study to evaluate the effectiveness of inactivated CoronaVac vaccine in preventing laboratory confirmed SARS-CoV-2 infection, among adults (aged ≥18 years) during January to June 2021. This test negative design was chosen due to several reasons. First, accessibility to individual data who were laboratory-tested for SARS-CoV-2 was feasible through the national surveillance system; second, it was cost-effective and allowed for faster results; and lastly, the design could better control for potential biases, including healthcare seeking behavior and access to COVID-19 testing [11].

### Data sources

We linked individual health electronic databases including SARS-CoV-2 reverse-transcription polymerase chain reaction (RT-PCR) test results from the national laboratory testing database (New All Record), COVID-19 vaccination registry, and hospitalization claims, managed by the Ministry of Health of Indonesia, using unique national citizen identification number. Data with foreign identification number and driving license number were excluded. We also checked the consistency in identification number, name, date of birth and sex between these databases. Confirmatory SARS-CoV-2 PCR test was performed on naso and/or oropharyngeal swab specimens in COVID-19 reference laboratories.[12]

### Selection of cases and matched controls

Cases were adults aged ≥18 years identified as those who had at least one COVID-19-like symptom (fever with body temperature documented at ≥38°C, chills, cough, shortness of breath, fatigue or malaise, sore throat, headache, runny nose, congestion, muscle aches, nausea or vomiting, diarrhea, abdominal pain, altered sense of smell or taste) and were tested RT-PCR positive for SARS-CoV-2, between January 13 to June 30, 2021; were not confirmed as positive for SARS-CoV-2 in the last 90 days before the index date; and those who did not receive any COVID-19 vaccines other than CoronaVac before sample collection. Controls were defined as those who had negative RT-PCR test results during the same period, irrespective for symptoms; and who did not receive any COVID-19 vaccines other than CoronaVac before sample collection. Cases and control were matched based on age, sex, district of residence and week of sample collection/testing.

### Vaccination status

Vaccination status of the subjects was determined by referring to linked individual vaccination record and was determined based on their SARS-CoV-2 test date. Fully vaccinated cases and controls were defined as having two doses of CoronaVac at least 14 days before the sample collection date. Adults who received no single dose of CoronaVac vaccine were defined as unvaccinated and all other adults who received one dose at least 14 days before sample collection date were defined as partially vaccinated.

### Statistical analysis

Unadjusted and adjusted odds of vaccination between test-positive and test-negative pairs were estimated using conditional logistic regression. Odds ratios (ORs) and 95% confidence interval (CI) of vaccination in cases and controls was compared. Adjusted ORs were estimated from models including age (by year, as continuous) and presence of comorbidities and prior SARS-CoV-2 infection. There was a considerable proportion of missing data for comorbidities (79%). Multiple imputation was performed with considering for covariates (age, sex, residence) and outcome. Vaccine effectiveness (VE) was calculated as (1-OR) x 100% and 95% CIs for vaccine effectiveness estimates as 100 × (1–upper or lower bounds of 95% CI for adjusted OR [13].

The effectiveness of CoronaVac against secondary outcomes (hospitalization and death due to COVID-19) were estimated using a multivariable logistic regression, accounting for covariates such as age, sex, district of residence, imputed presence of comorbidities and week of sample collection. Subgroup analyses were also performed to assess VE based on the age group. Sensitivity analysis was conducted on an unmatched population using a multivariable logistic regression model, adjusted for age, sex, district of residence, presence of comorbidities and week of testing. Statistical significance was set at p-value < 0.05. All statistical analyses were conducted using STATA software version 15.0 (Stata Cooperation, College Station, Texas, USA).

## Results

### Study population

Total 109,050 RT-PCR test results were retrieved during the January 13 to June 30, 2021 (Figure 1). Of which, 14,168 subjects were eligible for inclusion in the study. Of these, 2886 (20.4%) were tested positive for SARS-CoV-2 and 11,282 (79.6%) tested negative for SARS-CoV-2 infection. Of these, 5518 (39%) matched case-control pairs were eligible for analysis after matching by age, sex, geography (district of residence), and week of testing.

**Figure 1.**
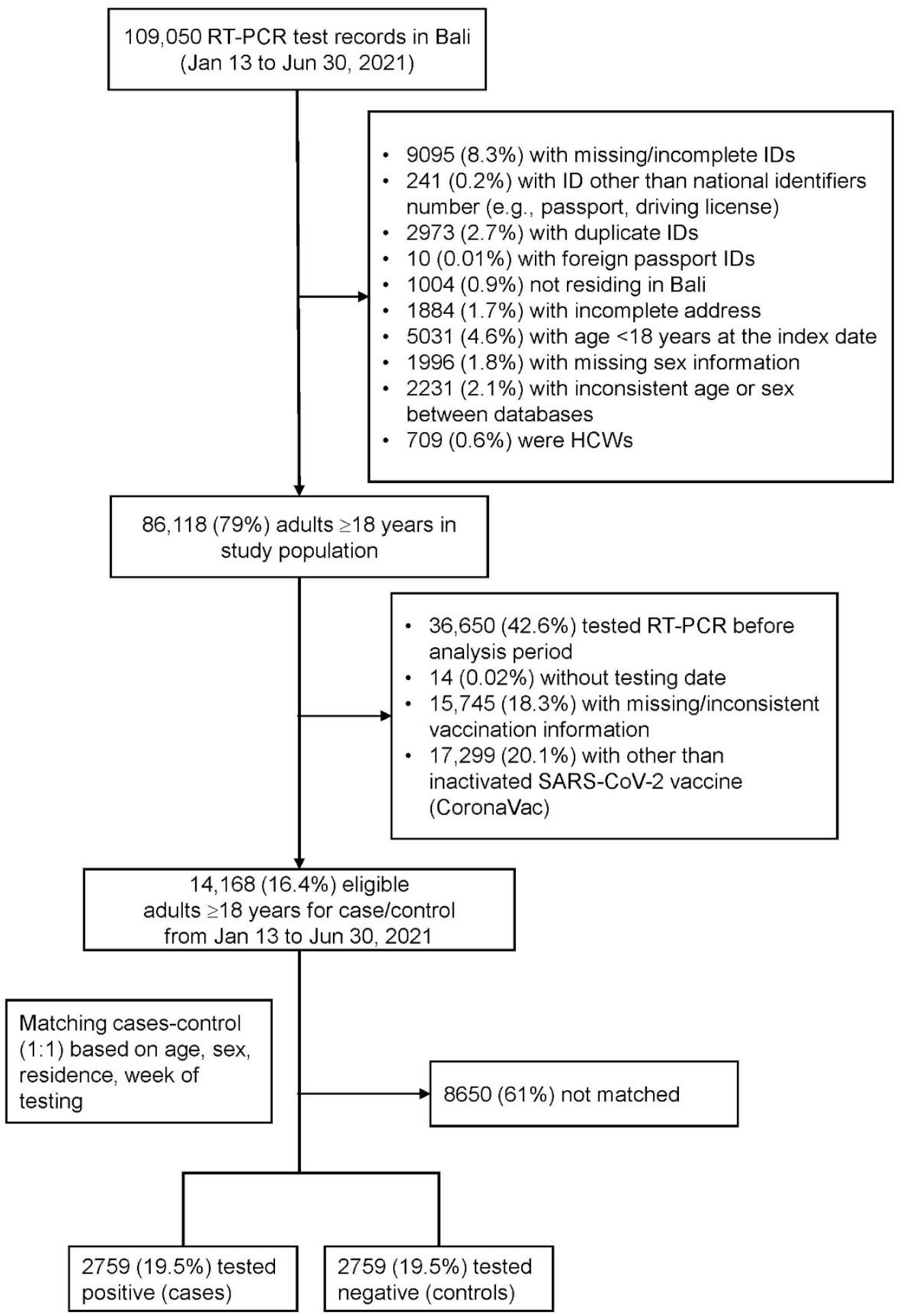
Flowchart for case and control selection

**Figure 2.**
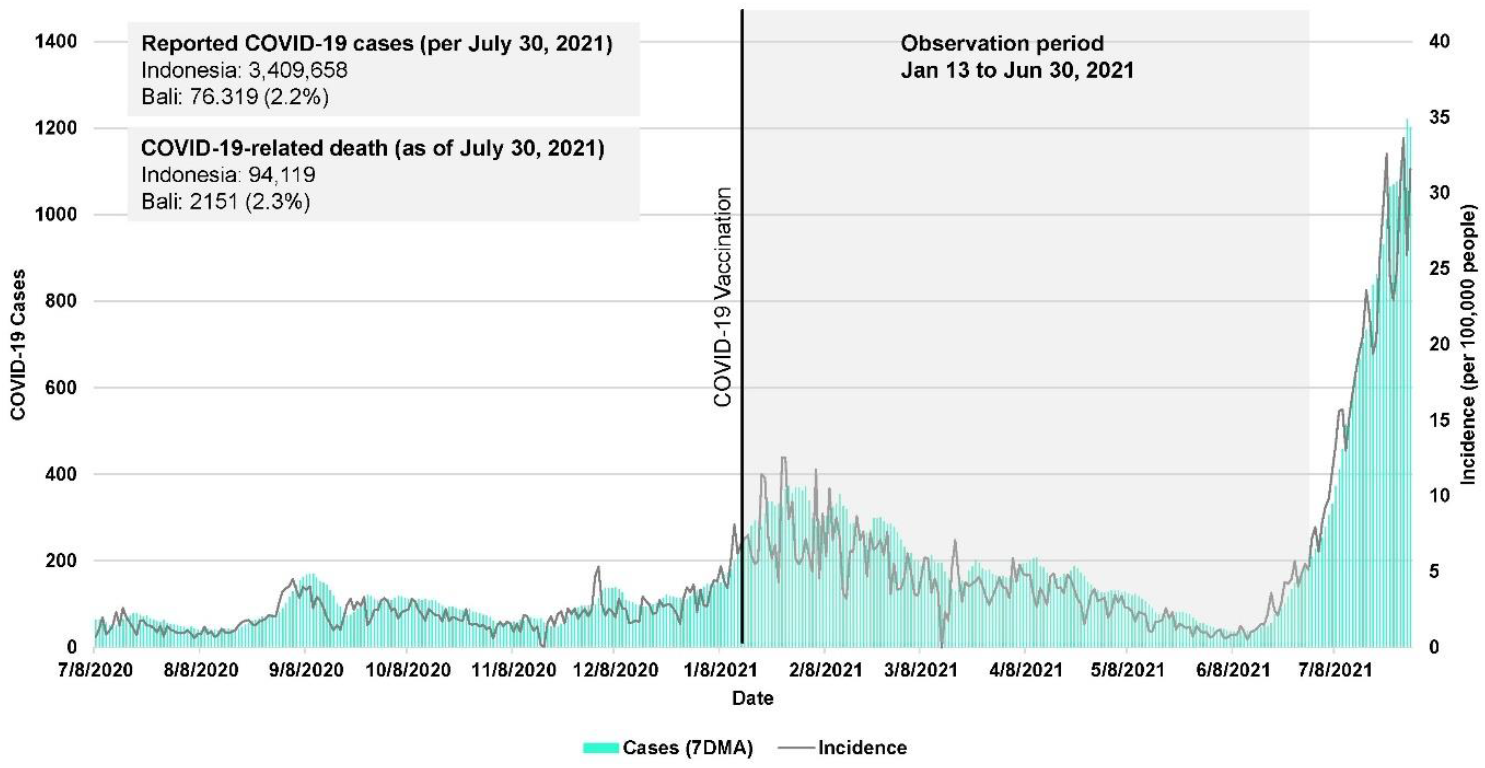
Trends in COVID-19 incidence in Bali over the period of study (Data source: Ministry of Health of Indonesia).

The characteristics of the study population and matched case-control pairs were presented in Table 1. The median age of the overall study sample (n=14,168) was 50 years (interquartile range, 31-65 years); 50.8% were aged ≥50 years. Thirty percent (n=4259) resided in Denpasar city. Thirty four percent (n=4863) had received full vaccination; 42.5% (n=6016) had not received vaccine. Those who receiving positive RT-PCR test results were appeared much older, males, had at least one comorbidity and unvaccinated.

**Table 1.** Characteristics of the adults aged ≥18 years eligible in the study and case-control pairs included in the analysis

| Characteristics | Total | Eligible study population |  | Matched pairs |  |
| --- | --- | --- | --- | --- | --- |
|  |  | Positive SARS-CoV-2 (n=2886) | Negative SARS-CoV-2 (n=11,282) | Cases (n=2759) | Controls (n=2759) |
| **Age, years, median (IQR)** | 50 (31-65) | 61 (45-69) | 46 (30-64) | 61 (45-69) | 60 (41-70) |
| **Age group, n (%)** |  |  |  |  |  |
| 18-49 y | 6969 (49.2) | 853 (29.6) | 6116 (54.2) | 847 (30.7) | 847 (30.7) |
| $\geq 50$ y | 7199 (50.8) | 2033 (70.4) | 5166 (45.8) | 1912 (69.3) | 1912 (69.3) |
| **Sex** |  |  |  |  |  |
| Male | 7676 (54.2) | 1730 (59.9) | 5946 (52.7) | 1648 (59.7) | 1648 (59.7) |
| Female | 6492 (45.8) | 1156 (40.1) | 5336 (47.3) | 1111 (40.3) | 1111 (40.3) |
| **District of residence, n (%)** |  |  |  |  |  |
| Jembrana | 564 (4) | 87 (3) | 477 (4.2) | 75 (2.7) | 75 (2.7) |
| Tabanan | 1500 (10.6) | 233 (8.1) | 1267 (11.2) | 233 (8.5) | 233 (8.5) |
| Badung | 2031 (14.3) | 474 (16.4) | 1557 (13.8) | 465 (16.7) | 456 (16.7) |
| Gianyar | 1573 (11.1) | 408 (14.1) | 1165 (10.3) | 363 (13.2) | 363 (13.2) |
| Klungkung | 554 (3.9) | 79 (2.7) | 475 (4.2) | 73 (2.7) | 73 (2.7) |
| Bangli | 803 (5.7) | 222 (7.7) | 581 (5.2) | 202 (7.3) | 202 (7.3) |
| Karangasem | 916 (6.5) | 144 (5) | 772 (6.8) | 138 (5) | 138 (5) |
| Buleleng | 1968 (13.9) | 353 (12.2) | 1615 (14.3) | 343 (12.4) | 343 (12.4) |
| Denpasar | 4259 (30.1) | 886 (30.7) | 3373 (29.9) | 867 (31.4) | 867 (31.4) |
| **Reported No. of comorbidities, n (%)** |  |  |  |  |  |
| No | 12,158 (85.8) | 1698 (58.8) | 10,460 (92.7) | 1636 (59.3) | 2473 (89.6) |
| Yes | 2010 (14.2) | 1188 (41.2) | 822 (7.3) | 1123 (40.7) | 286 (10.4) |
| **Vaccination status, n (%)** |  |  |  |  |  |
| Unvaccinated | 6016 (42.5) | 1867 (64.7) | 4149 (36.8) | 1771 (64.2) | 1469 (53.3) |
| 1st dose (0-13 days) | 1001 (7.1) | 222 (7.7) | 779 (6.9) | 216 (7.8) | 193 (7) |
| 1st dose ($\geq 14$ days) | 1408 (9.9) | 340 (11.8) | 1068 (9.5) | 319 (11.6) | 272 (9.9) |
| 2nd dose (0-13 days) | 880 (6.2) | 77 (2.7) | 803 (7.1) | 74 (2.7) | 158 (5.7) |
| 2nd dose ($\geq 14$ days) | 4863 (34.3) | 380 (13.2) | 4483 (39.7) | 379 (13.7) | 667 (24.2) |

Among the case subjects, about 40% of subjects having comorbidities. Based on vaccination status, 16,4% (n=453) had received two doses of CoronaVac vaccine. Nineteen percent (n=535) case subjects had received the first dose of CoronaVac vaccine and about two-third (n=1771) were unvaccinated. While among participants in the control group, half of the participants (n=1485/2759) were unvaccinated. Twenty four percent had received two doses of CoronaVac vaccine.

### Vaccine effectiveness

The unadjusted effectiveness of two doses of CoronaVac in preventing laboratory confirmed SARS-CoV-2 at least 14 days after administration was 73% (95% CI: 67-78) (Table 2). After adjusting for covariates, compared to unvaccinated adults, two dose of CoronaVac improved protection against SARS-CoV-2 infection (VE: 66.7%; 95% CI: 58.1-73.5) at ≥14 days after the second dose. Partial vaccination with CoronaVac was not significantly associated with a reduced risk of laboratory-confirmed SARS-CoV-2 infection (VE: 14.5%; 95% CI: -11 to 34.2) at 0-13 days after the first dose. The effectiveness of CoronaVac vaccine against COVID-19 infection was reduced with increasing age.

**Table 2.** CoronaVac vaccine effectiveness against laboratory-confirmed SARS-CoV-2 infection in adults aged ≥18 years in Bali, Indonesia, all age and stratified by age group

|  | Cases<br>(SARS-CoV-2<br>RT-PCR-positive) | Controls<br>(SARS-CoV-2<br>RT-PCR-negative) | Unadjusted<br>OR (95% CI) | Unadjusted<br>p-value | Adjusted OR*<br>(95% CI) | Adjusted<br>p-value | Adjusted VE<br>(95% CI) |
| --- | --- | --- | --- | --- | --- | --- | --- |
| Unvaccinated | 1771 (64.2) | 1469 (53.3) | Ref |  | Ref |  |  |
| 0-13 days after 1<sup>st</sup> dose | 216 (7.8) | 193 (7) | 0.77 (0.61-0.98) | 0.03 | 0.85 (0.66-1.11) | 0.239 | 14.5 (-11 – 34.2) |
| ≥14 days after 1<sup>st</sup> dose | 319 (11.6) | 272 (9.9) | 0.76 (0.62-0.93) | 0.007 | 0.89 (0.71-1.12) | 0.332 | 10.5 (-12 – 28.6) |
| 0-13 days after 2<sup>nd</sup> dose | 74 (2.7) | 158 (5.7) | 0.33 (0.24-0.45) | <0.0001 | 0.37 (0.26-0.52) | <0.0001 | 63.1 (48.5-73.6) |
| ≥14 days after 2<sup>nd</sup> dose | 379 (13.7) | 667 (24.2) | 0.27 (0.22-0.33) | <0.0001 | 0.33 (0.26-0.42) | <0.0001 | 66.7 (58.1-73.5) |
| **Age group: 18-49 years** |  |  |  |  |  |  |  |
| Unvaccinated | 247 (29.2) | 95 (11.2) | Ref |  | Ref |  |  |
| 0-13 days after 1<sup>st</sup> dose | 133 (15.7) | 122 (14.4) | 0.43 (0.29-0.62) | <0.0001 | 0.52 (0.35-0.77) | <0.0001 | 48.3 (22.6-65.5) |
| ≥14 days after 1<sup>st</sup> dose | 144 (17) | 131 (15.5) | 0.34 (0.23-0.51) | <0.0001 | 0.45 (0.30-0.68) | <0.0001 | 55.1 (32-70.4) |
| 0-13 days after 2<sup>nd</sup> dose | 39 (4.6) | 79 (9.3) | 0.20 (0.12-0.32) | <0.0001 | 0.22 (0.13-0.36) | <0.0001 | 78.2 (63.6-87) |
| ≥14 days after 2<sup>nd</sup> dose | 284 (33.5) | 420 (49.6) | 0.18 (0.12-0.25) | <0.0001 | 0.23 (0.16-0.33) | <0.0001 | 77 (66.6-84.2) |
| **Age group: ≥50 years** |  |  |  |  |  |  |  |
| Unvaccinated | 1524 (79.7) | 1374 (71.9) | Ref |  | Ref |  |  |
| 0-13 days after 1<sup>st</sup> dose | 83 (4.3) | 71 (3.7) | 1.05 (0.75-1.47) | 0.775 | 1.11 (0.74-1.64) | 0.617 | -10.6 (-64.3– 25.5) |
| ≥14 days after 1<sup>st</sup> dose | 175 (9.2) | 141 (7.4) | 1.02 (0.80-1.30) | 0.874 | 1.13 (0.85-1.51) | 0.396 | -13.2 (-50.6 – 14.9) |
| 0-13 days after 2<sup>nd</sup> dose | 35 (1.8) | 79 (4.1) | 0.38 (0.25-0.58) | <0.0001 | 0.42 (0.26-0.68) | <0.0001 | 58.1 (32.4-74) |
| ≥14 days after 2<sup>nd</sup> dose | 95 (4.9) | 247 (12.9) | 0.26 (0.19-0.35) | <0.0001 | 0.30 (0.21-0.42) | <0.0001 | 69.9 (57.7-78.7) |

Abbreviations: OR, odds ratio; VE, vaccine effectiveness
\*Adjusted for age (as continuous), presence of comorbidities and prior SARS-CoV-2 infection
† Vaccine effectiveness is calculated as (1- odds ratio) x 100%

The adjusted VE in preventing COVID-19-related hospitalization and death was 71.1% (95% CI: 62.9-77.6) and 87.4% (95% CI: 65.1-95.4), respectively, at 14 days after the second dose (Table 3). Partial vaccination was less protective against COVID-19-related hospitalization and death among people aged 18 and older. Vaccine effectiveness against hospitalization was higher 79.4% (95% CI: 70-85.9) in group of people aged ≥50 years relative to people aged 18-49 years (65.8%; 95% CI: 49.1-77). Death due to COVID-19 was not significantly associated with full vaccination in people aged 18-49 years (VE: 65.7%, 95% CI: -99.5 to 94.1). This result could be happened due to an effect of small sample size and small number of events (mortality). Among older population, vaccine effectiveness was 90.6% (95% CI:61.5-97.7) in preventing death due to COVID-19.

**Table 3.** CoronaVac vaccine effectiveness in preventing hospitalization and COVID-19-related death in adults aged ≥18 years in Bali, Indonesia, all ages and stratified by age group

|  | Hospitalization |  |  |  | Death due to COVID-19 |  |  |  |
| --- | --- | --- | --- | --- | --- | --- | --- | --- |
|  | n/N | Adjusted OR<br>(95% CI) | Adjusted<br>p-value | VE*<br>% (95%CI) | n/N | Adjusted OR<br>(95% CI) | Adjusted<br>p-value | VE*<br>% (95%CI) |
| Unvaccinated | 1932/3240 | Ref |  |  | 450/3240 | Ref |  |  |
| 0-13 days after 1<sup>st</sup> dose | 124/409 | 0.72 (0.54-0.95) | 0.019 | 28.3 (5.3-45.7) | 10/409 | 0.46 (0.24-0.90) | 0.023 | 53.9 (10-76.3) |
| $\geq 14$ days after 1<sup>st</sup> dose | 218/591 | 0.66 (0.52-0.84) | 0.001 | 34.1 (16.4-48.1) | 15/591 | 0.41 (0.24-0.72) | 0.002 | 58.6 (28.3-76.1) |
| 0-13 days after 2<sup>nd</sup> dose | 50/232 | 0.39 (0.26-0.57) | <0.0001 | 61.2 (42.6-73.7) | 3/232 | 0.24 (0.07-0.77) | 0.017 | 76 (22.9-92.5) |
| $\geq 14$ days after 2<sup>nd</sup> dose | 166/1048 | 0.29 (0.22-0.37) | <0.0001 | 71.1 (62.9-77.6) | 4/1046 | 0.13 (0.05-0.35) | <0.0001 | 87.4 (65.1-95.4) |
| **Age group: 18-49 years** |  |  |  |  |  |  |  |  |
| Unvaccinated | 123/342 |  |  | Ref | 11/342 |  |  | Ref |
| 0-13 days after 1<sup>st</sup> dose | 53/255 | 0.66 (0.43-1.02) | 0.062 | 33.7 (-2.1-56.9) | 0/255 | 1 | - | NA** |
| $\geq 14$ days after 1<sup>st</sup> dose | 65/275 | 0.67 (0.44-1.02) | 0.064 | 32.6 (-2.3-55.6) | 1/275 | 0.22 (0.02-2.05) | 0.184 | 77.9 (-105.4-97.6) |
| 0-13 days after 2<sup>nd</sup> dose | 16/118 | 0.33 (0.17-0.65) | 0.001 | 66.9 (35-83.2) | 1/118 | 0.30 (0.03-2.97) | 0.303 | 70 (-196.5-97) |
| $\geq 14$ days after 2<sup>nd</sup> dose | 99/704 | 0.35 (0.23-0.52) | <0.0001 | 65.1 (48-76.5) | 2/704 | 0.45 (0.07-2.80) | 0.390 | 55.3 (-180.2-92.9) |
| **Age group: $\geq 50$ years** |  |  |  |  |  |  |  |  |
| Unvaccinated | 1809/2898 |  |  | Ref | 439/2898 |  |  | Ref |
| 0-13 days after 1<sup>st</sup> dose | 71/154 | 0.86 (0.58-1.27) | 0.446 | 14.1 (-27.1-42) | 10/154 | 0.65 (0.33-1.28) | 0.213 | 34.9 (-28-66.9) |
| $\geq 14$ days after 1<sup>st</sup> dose | 153/316 | 0.67 (0.50-0.90) | 0.009 | 32.9 (9.5-50.2) | 14/316 | 0.45 (0.25-0.79) | 0.005 | 55.2 (21.1-74.6) |
| 0-13 days after 2<sup>nd</sup> dose | 34/114 | 0.44 (0.27-0.72) | 0.001 | 55.9 (28-72.9) | 2/114 | 0.20 (0.05-0.82) | 0.025 | 80.2 (18.1-95.2) |
| $\geq 14$ days after 2<sup>nd</sup> dose | 67/342 | 0.21 (0.15-0.30) | <0.0001 | 79 (69.7-85.5) | 2/342 | 0.09 (0.02-0.37) | 0.001 | 90.9 (62.6-97.8) |

Abbreviations: OR, odds ratio; VE, vaccine effectiveness; CI, confidence interval
\*Adjusted for age (as continuous), sex, presence of comorbidities and prior SARS-CoV-2 infection, week of testing, district of residence. OR is estimated using multivariable regression model.
\*\*The CI could not be estimated owing to zero/few events in the group.

To address potential issues related to the small sample size and rare event or outcome, we conducted a sensitivity analysis using unmatched population (n=14,168) (Supplementary Table S1 - Table S3). Overall, after adjusting all covariates, the vaccine effectiveness was 66.7% (95%CI: 61-71.5) in preventing laboratory confirmed SARS-CoV-2 infection at 14 days or more after the second dose administration. The effectiveness was relatively higher in adults aged 18-49 years (VE: 76.9; 95%CI: 77.3-86.6) compared to that older participants (VE: 69.4%, 95%CI: 60.8-76.2), at 14 days after the second dose.

The effectiveness against hospitalization and death related to COVID-19 was 77.7% (95%CI: 73.2-81.5) and 93.9% (95%CI: 85-97.6%), respectively, at 14 days after second dose. The effectiveness against both hospitalization and death due to COVID-19 was higher in that population aged 50 years or older (82.6% and 94.4%, respectively) compared to group of people aged 18-49 years (71.8% and 88.1%, respectively).

## Discussion

High proportion of inactivated vaccine of CoronaVac have been used in national COVID-19 vaccination program in Indonesia. So far, this is the first health record-based observational study that provide real-world evidence on the effectiveness of COVID-19 vaccine against laboratory-confirmed SARS-CoV-2 infection, hospitalization, and death in Indonesian population. The study showed complete regimen of CoronaVac vaccine at least 14 days after the second dose had an effectiveness of 66.7% (95% CI: 58.1-73.5) against COVID-19 infection among Balinese adults aged ≥18 years. This finding is consistent with the results of phase III trial [5]. Yet, the effectiveness against infection was reduced by age. The effectiveness against infection was slightly lower in elderly aged 50 years and above (69.9%; 95% CI: 57.7-78.7) compared to younger adults (18-49 years) (77%, 95% CI: 66.6-84.2). This could be associated with the difference in immune response. It has been observed in older people that the antibody titre was rapidly degraded, the immune response was delay and the peak of neutralizing antibodies was relatively lower than younger populations [14]. These findings highlight the necessity for elderly-specific immunization approaches (e.g., vaccine formulations and administration).

Our real-world evidence on the effectiveness of CoronaVac against laboratory confirmed SARS-CoV-2 infection was consistent with that randomized controlled trials of vaccine efficacy in Indonesia (65.3%; 95%CI: 20-85.1) [2]. Yet, it was lower than that of vaccine efficacy reported in Turkey (83.5%; 95% CI: 65.4-92.1) and higher than that efficacy reported from Brazil (51%; 95%CI: 36-62) at 14 days after the second vaccination [3, 15]. Chilean cohort VE study involving general population aged ≥16 years old also demonstrated the VE was 65.9% (95%CI: 65.2%-66.6%) against COVID-19 infection. While a TND case control study in Brazil demonstrated that two dose regimens of CoronaVac was effective in reducing risk of symptomatic COVID-19 (46.8%; 95%CI: 38.7% to 53.8%) at ≥14 days after the second dose among older population aged ≥70 years [7]. Another inactivated vaccine platform, BBV152 (a whole-virion inactivated vaccine manufactured by Bharat Biotech, Covaxin) which is used in India, has also been reported to be effective against COVID-19. A study reported that the effectiveness of against symptomatic COVID-19 was 50% (95% CI: 33–62) [16]. Compared to the effectiveness of CoronaVac vaccine, BBV152 effectiveness is slightly lower. The difference in the amount of transmission and the circulating SARS-CoV-2 variant are likely to have an impact on the results.

Our study showed that full vaccination of CoronaVac vaccine was also effective in preventing hospitalization and COVID-19-associated death. We estimated that CoronaVac vaccine was 71.1% (95% CI: 62.9-77.6) for the prevention of hospitalization and 87.4% (95%CI: 65.1-95.4) for the prevention of COVID-19-related death at least 14 days after the second dose. While partial vaccination provided less protection against hospitalization and death due to COVID-19. None has reported CoronaVac VE estimates against these two outcomes in Indonesia. Our estimates, however, are dissimilar to that estimate reported elsewhere. A cohort study in Chile among adults aged ≥16 years showed that CoronaVac vaccine was 87.5% (95% CI, 86.7-88.2) effective against hospitalization and 86.3% (95% CI, 84.5-87.9) effective against COVID-19– related death [6, 17]. While a TND study in Brazil among elderly aged 70 years or older, CoronaVac was effective against COVID-19-related hospitalization (55.5%; 95%CI: 46.5-62.9) and death (61.2%, 95%CI: 48.9-70.5) at ≥14 days after the second dose, respectively [7]. A study on another inactivated vaccine showed that the effectiveness of full vaccination of Sinopharm BBIBP-CorV was 79.8% (95% CI: 78–81.4) in preventing hospital admission and 97.1% (83–99.9) in preventing death among Abu Dhabi residents aged 15 years and older [18]; which is slightly higher than our estimates on the effectiveness of CoronaVac. This discrepancy might be due to several factors including variation in study population and design (e.g., targeted age group, sample size) and difference in level of transmission or epidemiological setting (e.g., a TND study in Brazil was conducted when P.1 or Gamma variant was predominant).

### Our adjusted effectiveness

The strengths of this study are that: i) we used comprehensive health administrative database including individual laboratory test results, vaccination data and hospitalizations which include information on date of sample collection, type of vaccine, date of vaccination, date of hospital admission and underlying conditions; (ii) the subjects were chosen based on a standard laboratory confirmation for SARS-CoV-2 results. Thus, biases due to misclassification from infection or vaccination status were implausible. These findings are important to fill the gap in our knowledge regarding the performance of inactivated CoronaVac vaccine in the real-world; (iii) our analysis have sufficient of power (99.3%) as our matched case-control study involved large number of samples (2759 cases) with known vaccine effectiveness of 65% against symptomatic SARS-CoV-2 ≥14 days after the second dose.

However, our study has limitations. First, while we attempted to make a robust approach by adjusting for number of potential confounding factors, including age, sex, geography, week of testing (sample collection) and presence of comorbidities, there might be still unmeasured confounding factors that biased the VE estimates (e.g., behavioral risk factors, type of variants). Second, we did not have a detailed information on symptoms and date of onset in the record so that we could not identified and specifically examine the effectiveness against symptomatic cases. Third, as the data for date on onset were not available in the records, we used date of sample collection instead. There might be false-negative results included in the study so that this could affect our estimate. Lastly, this study was retrieved data from January to June 2021, which was just before the peak of Delta variants (B.1.617.2) outbreaks. Hence, the VE estimates presented here is likely reflected vaccine performance before Delta and Omicron becoming predominant variant in the community. VE of CoronaVac against Delta and Omicron-related infection and severity might be different. Studies elsewhere have reported the effects of Delta variants on the COVID-19 vaccines performance [19, 20]. Further research is required to evaluate the effectiveness of CoronaVac against the circulating VOCs.

## Conclusions

The two-dose regimens of inactivated CoronaVac vaccine was effective in preventing COVID-19 infection, hospitalizations for COVID-19 and COVID-19-related death in adults aged 18 years or older. Full vaccination was significantly reduced the risk of severe outcomes especially among 50 years or older.

## Data Availability

The datasets used and/or analyzed during the current study are protected by the Ministry of Health of Indonesia and are unsuitable for public sharing. Interested parties can apply for the data by contacting the data center of the Ministry of Health of Indonesia. All data generated or analyzed during this study are included in this published article (and its Supplementary information files). Aggregate data on vaccination and COVID-19 incidence are publicly available at https://vaksin.kemkes.go.id/#/vaccines

## Author contributions

Study conceptualization: AS, PWD, RR (Ririn Ramadhany), MH. Methodology: AS, RR (Ririn Ramadhany), MH, PWD. Formal analysis: AS, RA, EHF, PWD, MH. Project administration: RA, RR (Rustika Rustika), KLT, TT. Data collection: AS, RA, EHF, AA, ILI, PWD. Supervision: PWD, RR (Ririn Ramadhany). Validation: PWD, RR (Rustika Rustika), KLT, TT. Funding: AS, TT, MH. Writing (original draft): AS, PWD. Writing (review and editing): all authors. All authors have read and approved the manuscript.

## Ethic statement

The protocol of the study was approved by the Health Research Ethics Committee, National Institute of Health Research and Development, Ministry of Health, Indonesia (LB. 02.01/2/KE.533/2021). The study was considered exempt from informed consent. No human health risks were identified.

## Acknowledgment

We would like to thank Dr. Anas Ma’ruf and team at the Center of Health Data and Information of Ministry of Health Indonesia, and Mr. Made Darmawan and Mr. Akbar Antony Siregar at the National COVID-19 Control and Economic Recovery Committee (KPC-PEN) for their assistance and support in collating the data. We would also like to thank the Ketut Suarjaya and I Wayan Widia at Bali Provincial Health Office for their support in the implementation of this study and permission to access COVID-19 surveillance data.

